# Development and External Validation of a Diagnostic Model for Coronary Microvascular Obstruction

**DOI:** 10.1101/2020.06.27.20141416

**Authors:** Yong Li, Shuzheng Lyu

**Affiliations:** Emergency and Critical Care Center, Beijing Anzhen Hospital, Capital Medical University, Beijing 100029, China; Department of Cardiology, Beijing Anzhen Hospital, Capital Medical University, Beijing 100029, China

**Keywords:** coronary disease, ST elevation myocardial infarction, no-reflow phenomenon, percutaneous coronary intervention, nomogram

## Abstract

**Background:** Prevention of coronary microvascular obstruction /no-reflow phenomenon(CMVO/NR) is a crucial step in improving prognosis of patients with acute ST segment elevation myocardial infarction (STEMI)during primary percutaneous coronary intervention (PPCI). We wanted to develop and externally validate a diagnostic model of CMVO/NR in patients with acute STEMI underwent PPCI.

**Methods:** Design: Multivariable logistic regression of a cohort of acute STEMI patients. Setting: Emergency department ward of a university hospital. Participants: Diagnostic model development: Totally 1232 acute STEMI patients who were consecutively treated with PPCI from November 2007 to December 2013. External validation: Totally 1301 acute STEMI patients who were treated with PPCI from January 2014 to June 2018. Outcomes: CMVO/NR during PPCI.

**Results:** 147(11.9%)patients presented CMVO/NR in the development dataset and 120(9.2%) patients presented CMVO/NR in the validation dataset. The strongest predictors of CMVO/NR were age, periprocedural bradycardia, using thrombus aspiration devices during procedure and total occlusion of culprit vessel. We developed a diagnostic model of CMVO/NR.The area under the receiver operating characteristic curve (AUC) was 0.6833 in the development set.We constructed a nomogram using the development database.The AUC was 0.6547 in the validation set. Discrimination, calibration, and decision curve analysis were satisfactory.

**Conclusions:** We developed and externally validated a diagnostic model of CMVO/NR during PPCI.

We registered this study with WHO International Clinical Trials Registry Platform on 16 May 2019. Registration number: ChiCTR1900023213. http://www.chictr.org.cn/edit.aspx?pid=39057&htm=4.

## Introduction

Although primary percutaneous coronary intervention (PPCI) is the most beneficial reperfusion strategy available to patients with acute ST segment elevation myocardial infarction (STEMI), PPCI may not restore the best myocardium in myocardial tissue reperfusion.A failure at the microvascular level known as coronary microvascular obstruction/no-reflow phenomenon(CMVO/NR). ^[1-3]^ Coronary angiography performed during PPCI at the acute phase of myocardial infarction is the first imaging technique to identify the CMVO/NR in humans. ^[4]^ CMVO/NR is defined as Thrombolysis In Myocardial Infarction flow grade (TIMI) < 3. ^[2,3]^ CMVO/NR can result in poor healing of the infarct and adverse left ventricular remodeling, increasing the risk for major adverse cardiac events.^[2]^Prevention of CMVO/NR is therefore a crucial step. We developed and externally validated a diagnostic model of CMVO/NR. We constructed a nomograms using the development database.

The aim of this study was 4-fold: (1) to identify predictive factors; (2) to develop a diagnostic model; (3) to create a nomogram and (4) to externally validate diagnostic model.

## Methods

We followed the methods of Li et al. 2019 ^[5]^and Li. 2020 ^[6]^.

We followed the methods of Transparent Reporting of a multivariable prediction model for Individual Prognosis Or Diagnosis (TRIPOD). ^[7]^We used type 2b of prediction model studies covered by the TRIPOD statement. We split the data nonrandomly by time into 2 groups: one to develop the prediction model and one to evaluate its predictive performance. ^[7]^Type 2b was referred to as “external validation studies”. ^[7]^

The derivation cohort was 1232 patients with acute STEMI presenting within 12 hours from the symptom onset who were consecutively treated with PPCI between November 2007 and December 2013 in Beijing Anzhen Hospital, Capital Medical University.

The validation cohort was 1301 patients we recruited on the same basis between January 2014 and June 2018 in Beijing Anzhen Hospital, Capital Medical University.

Inclusion criteria:1. patients with acute STEMI presenting within 12 hours from the symptom onset who were treated with PPCI; 2. age of more than 18 years and less than 80-year-old male and non-pregnant women. ^[5]^We established the diagnosis of acute myocardial infarction (AMI) and STEMI base on the fourth universal definition of myocardial infarction(MI). ^[8]^

Exclusion criteria: 1. patients received thrombolytic; 2. patients received bivalirudin.

Prior to emergency angiography, all patients received 300 mg of aspirin, 300 to 600 mg of clopidogrel or 180 mg of ticagrelor and unfractionated heparin(patients received bivalirudin were not included). ^[5]^

It was a retrospective analysis and informed consent was waived by Ethics Committee of Beijing Anzhen Hospital Capital Medical University.

Outcome of interest was CMVO/NR. CMVO/NR was defined as TIMI <3 after successful mechanical opening of the infarct-related arterie with a guide wire, a balloon, or a stent, not the end of the PPCI process. ^[2,9,10]^The presence or absence of CMVO/NR was decided blinded to the predictor variables and based on consensus.

We selected 11 predictors based on clinical relevance and the results of the pre-experiment cohort. The potential candidate variables were age,sex,medical history such as hypertension,diabetes, angina, myocardial infarction, and percutaneous coronary intervention(PCI), culprit vessel site, total occlusion of culprit vessel, the time between myocardial infarction and PPCI, using thrombus aspiration devices during procedure, periprocedural bradycardia(pre-procedural heart rate was greater than or equal to 50 times / min, intra-procedural heart rate was less than 50 times / min persistent or transient), ^[11]^ and intra-procedural hypotension(pre-procedural systolic blood pressure was greater than 90/60mmHg, intra-procedural systolic blood pressure was less than or equal to 90 mmHg persistent or transient). ^[12]^

Pre-procedural blood pressure, pre-procedural heart rate, age, sex, medical history such as hypertension, diabetes, angina, myocardial infarction, and PCI was based on the medical record. CMVO/NR, culprit vessel site, total occlusion of culprit vessel, periprocedural bradycardia, and intra-procedural hypotension was based on the procedure record.

Some people suggested that each candidate variable must have at least 10 events for model derivation and at least 100 events for model validation. ^[7]^ Our number of samples and events exceeded expectations, so it was expected to provide very reliable estimates. ^[7]^

To ensure reliability of data, we excluded patients who had missing information on key predictors: age, periprocedural bradycardia, using thrombus aspiration devices during procedure, and total occlusion of culprit vessel.

## Statistical Analysis

We presented data as mean ±SD or n(%).We kept all continuous data as continuous and retained on the original scale. We used univariable and multivariable logistic regression models to identify the correlates of CMVO/NR during PPCI. We entered all variables of Tables 1 into the univariable logistic regression. We constructed a multivariable logistic regression model using the backward variable selection method, based on the variables that resulted significant from univariable logistic regression. ^[5]^

**Table 1.**
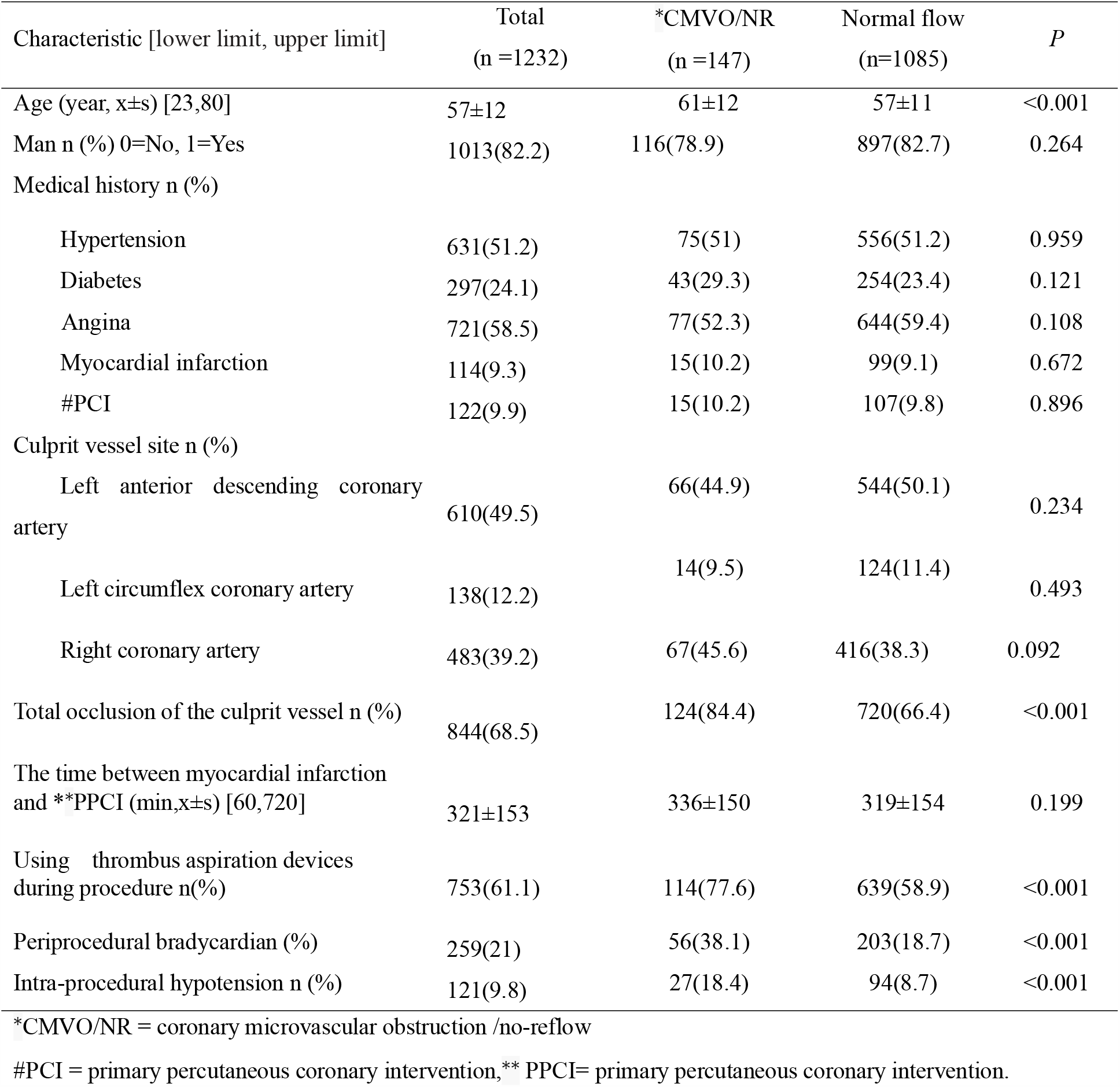
Demographic, clinical, and angiographic characteristics of patients with CMVO/NR and normal coronary flow during PPCI in the development data sets.

We used the Akanke information criterion (AIC)and Bayesian information criterion(BIC)to select predictors; It accounts for model fit while penalizing for the number of parameters being estimated and corresponds to using α =0.157. ^[7]^

We assessed the predictive performance of the diagnostic model in the validation data sets by examining measures of discrimination, calibration and decision curve analysis (DCA).

Statistical analyses were performed with STATA version 15.1 (StataCorp, College Station, TX) , R version 3.5.1(R Development Core Team; http://www.r-project.org) and the RMS package developed by Harrell(Harrell et al).All tests were two-sided and a P value <0.05 was considered statistically significant.

All methods were carried out in accordance with relevant guidelines and regulations.

## Results

We drew a flow diagrams (Figure 1).

**Figure 1.**
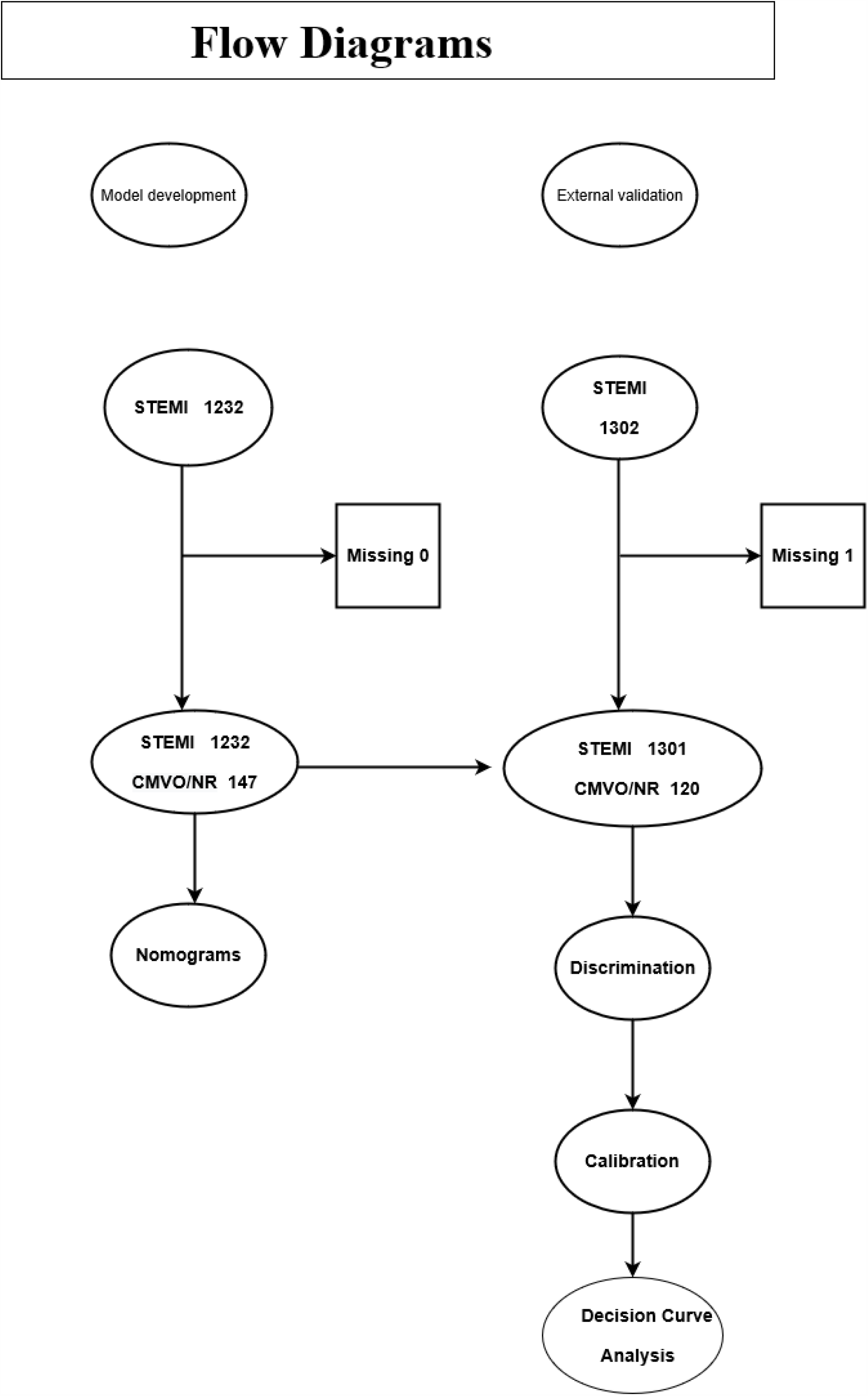
Flow diagrams.

Totally 1085 patients had a TIMI flow grade 3 (group with normal epicardial flow) and 147 patients had a TIMI flow grade 0∼ 2(group with CMVO/NR) in the development data set. Baseline characteristics of the patients was shown in Table 1. Eight variables (total occlusion of culprit vessel, age, history of angina, history of diabetes, periprocedural bradycardia, intra-procedural hypotension, using thrombus aspiration devices during procedure, and the culprit vessel was right coronary artery(RCA)were significant differences in the two groups of patients(p < 0. 157). After application of backward variable selection method, AIC and BIC, four variables (age, periprocedural bradycardia, using thrombus aspiration devices during procedure, and total occlusion of culprit vessel) remained as significant independent predictors of CMVO/NR during PPCI. Results were shown in Table 2 and Table3.

**Table 2.**
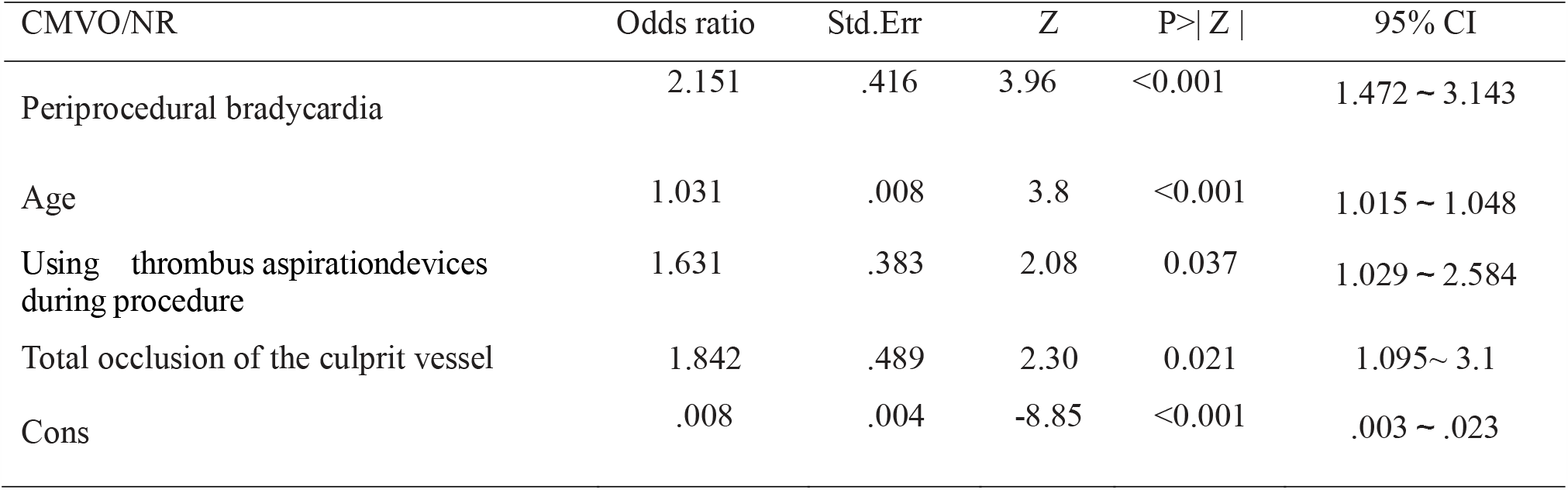
Predictor of CMVO/NR obtained from multivariable logistic regression models (odds ratio) in the development data sets

| CMVO/NR | Odds ratio | Std.Err | Z | P>\| Z \| | 95% CI |
| --- | --- | --- | --- | --- | --- |
| Periprocedural bradycardia | 2.151 | .416 | 3.96 | <0.001 | 1.472 ~ 3.143 |
| Age | 1.031 | .008 | 3.8 | <0.001 | 1.015 ~ 1.048 |
| Using thrombus aspiration devices during procedure | 1.631 | .383 | 2.08 | 0.037 | 1.029 ~ 2.584 |
| Total occlusion of the culprit vessel | 1.842 | .489 | 2.30 | 0.021 | 1.095~ 3.1 |
| Cons | .008 | .004 | -8.85 | <0.001 | .003 ~ .023 |

**Table 3.** Predictor of CMVO/NR obtained from multivariable logistic regression models (Coef) in the development data sets

| CMVO/NR | Coef | Std.Err | Z | P>\| Z \| | 95% CI |
| --- | --- | --- | --- | --- | --- |
| Periprocedural bradycardia | .766 | .194 | 3.96 | <0.001 | .387 ~ 1.145 |
| Age | .031 | .008 | 3.8 | <0.001 | .015 ~ .047 |
| Using thrombus aspiration devices during procedure | .489 | .235 | 2.08 | 0.037 | .029 ~ .949 |
| Total occlusion of the culprit vessel | .611 | .266 | 2.3 | 0.021 | .091~1.132 |
| Cons | -4.831 | .546 | -8.85 | <0.001 | -5.902 ~ -3.761 |

According to the above risk factors,we can calculate the predicted probability of CMVO/NR using the following formula: P = 1 /(1 + exp(-(-4.831+.766×PB+.031× AGE(year)+.611×TOCV+.489×TA))). PB=periprocedural bradycardia (0=No, 1=Yes), TOCV=total occlusion of the culprit vessel(0=No, 1=Yes), TA = using thrombus aspiration devices during procedure (0=No, 1=Yes). The receiver operating characteristic curve (ROC) curve was drawn. The area under the receiver operating characteristic curve (AUC) was 0.6833±0.023, 95% CI= 0.639∼0.728. We constructed the nomogram (Figure 2)using the development database based on the four independent prognostic markers: age, total occlusion of culprit vessel, using thrombus aspiration devices during procedure,and periprocedural bradycardia.

**Figure 2.**
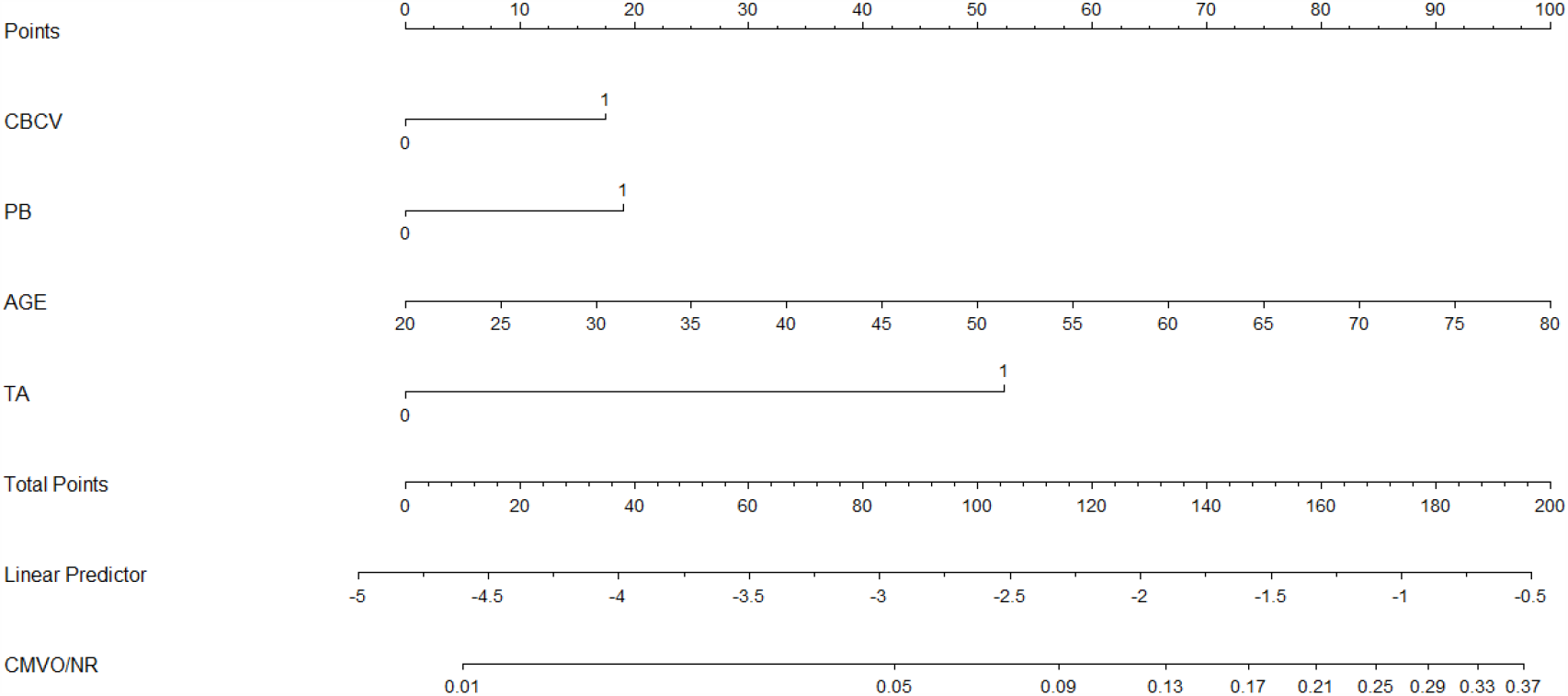
A nomograms for predicting CMVO/NR during PPCI in patients with acute STEMI AGE=Age(year); CMVO/NR = coronary microvascular obstruction /no-reflow phenomenon; CBCV=total occlusion of the culprit vessel(0=No, 1=Yes); PB= periprocedural bradycardia (0=No, 1=Yes);TA = using thrombus aspiration devices during procedure (0=No, 1=Yes).

Totally 1301 acute STEMI patients who were treated with PPCI from January 2014 to June 2018 in Beijing Anzhen Hospital, Capital Medical University.Baseline characteristics of the patients was shown in Table 4.

**Table 4.** Demographic, clinical, and angiographic characteristics of patients with CMVO/NR and normal coronary flow during PPCI in the validation data sets.

| Characteristic [lower limit, upper limit] | Total<br>(n =1301) | *CMVO/NR<br>(n =120) | Normal Flow<br>(n=1181) | *P* |
| --- | --- | --- | --- | --- |
| Age (year, x±s) [25,80] | 56±12 | 59±10 | 56±11 | 0.003 |
| Man n (%) 0=No, 1=Yes | 1097(84.3) | 93(77.5) | 1004(85) | 0.033 |
| Medical historyn (%) |  |  |  |  |
| Hypertension | 694(53.3) | 73(60.8) | 621(52.6) | 0.085 |
| Diabetes | 354(27.2) | 35(29.2) | 319(27) | 0.613 |
| Angina | 439(33.7) | 37(30.8) | 402(34) | 0.480 |
| Myocardial infarction | 91(7) | 10(8.3) | 81(6.9) | 0.547 |
| #PCI | 127(9.7) | 18(15) | 109(9.2) | 0.045 |
| Culprit vessel site n (%) |  |  |  |  |
| Left anterior descending coronary artery | 599(46) | 56(46.7) | 543(46) | 0.885 |
| Left circumflex coronary artery | 164(12.6) | 8(6.7) | 156(13.2) | 0.044 |
| Right coronary artery | 541(41.6) | 56(46.7) | 485(41.1) | 0.236 |
| The time between myocardial infarction and **PPCI (min,x±s) [60,720] | 346±156 | 365±151 | 344±156 | 0.159 |
| Using thrombus aspiration devices during procedure n(%) | 806(62) | 93(77.5) | 713(60.4) | <0.001 |
| Total occlusion of the culprit vessel n (%) | 896(68.9) | 98(81.7) | 798(67.6) | 0.002 |
| Periprocedural bradycardian (%) | 175(13.5) | 26(21.7) | 149(12.6) | 0.006 |
| Intra-procedural hypotension n (%) | 123(9.5) | 19(15.8) | 104(8.8) | 0.014 |

Abbreviations as in Table 1.

We can calculate the predicted probability of CMVO/NR using the following formula:

P = 1 /(1 + exp(-(-4.831+.766×PB+.031×AGE(year)+.611×TOCV+.489×TA))).

PB=periprocedural bradycardia(0=No, 1=Yes), TOCV=total occlusion of the culprit vessel(0=No, 1=Yes), TA = using thrombus aspiration devices during procedure(0=No, 1=Yes). The ROC curve was drawn.

AUC was 0.6547±0.025,95% CI= 0.605∼0.704.

We drew a calibration plot(Figure 3) with distribution of the predicted probabilities for individuals with and without the CMVO/NR in the validation data sets.Hosmer-Lemeshow chi2(10) = 16.26, Prob > chi2 = 0.0925>0.05.

**Figure 3.**
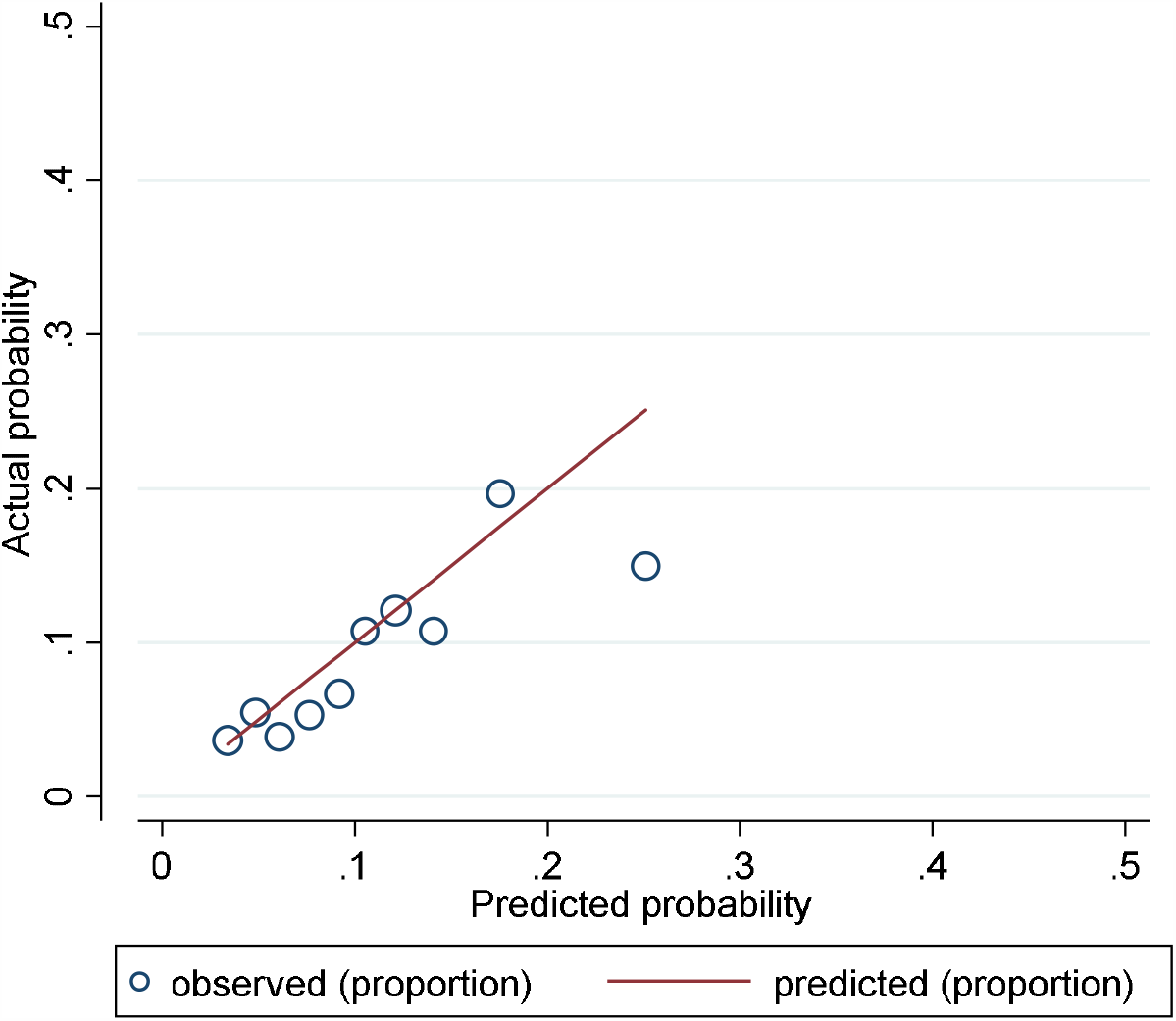
A calibration plot with distribution of the predicted probabilities for individuals with and without the CMVO/NR in the validation data sets.

Hosmer-Lemeshow chi2(10) = 16.26,Prob > chi2 = 0.0925>0.05. Brier score = 0.0997<0.25.

DCA(Figure 4)in the validation data sets.

**Figure 4.**
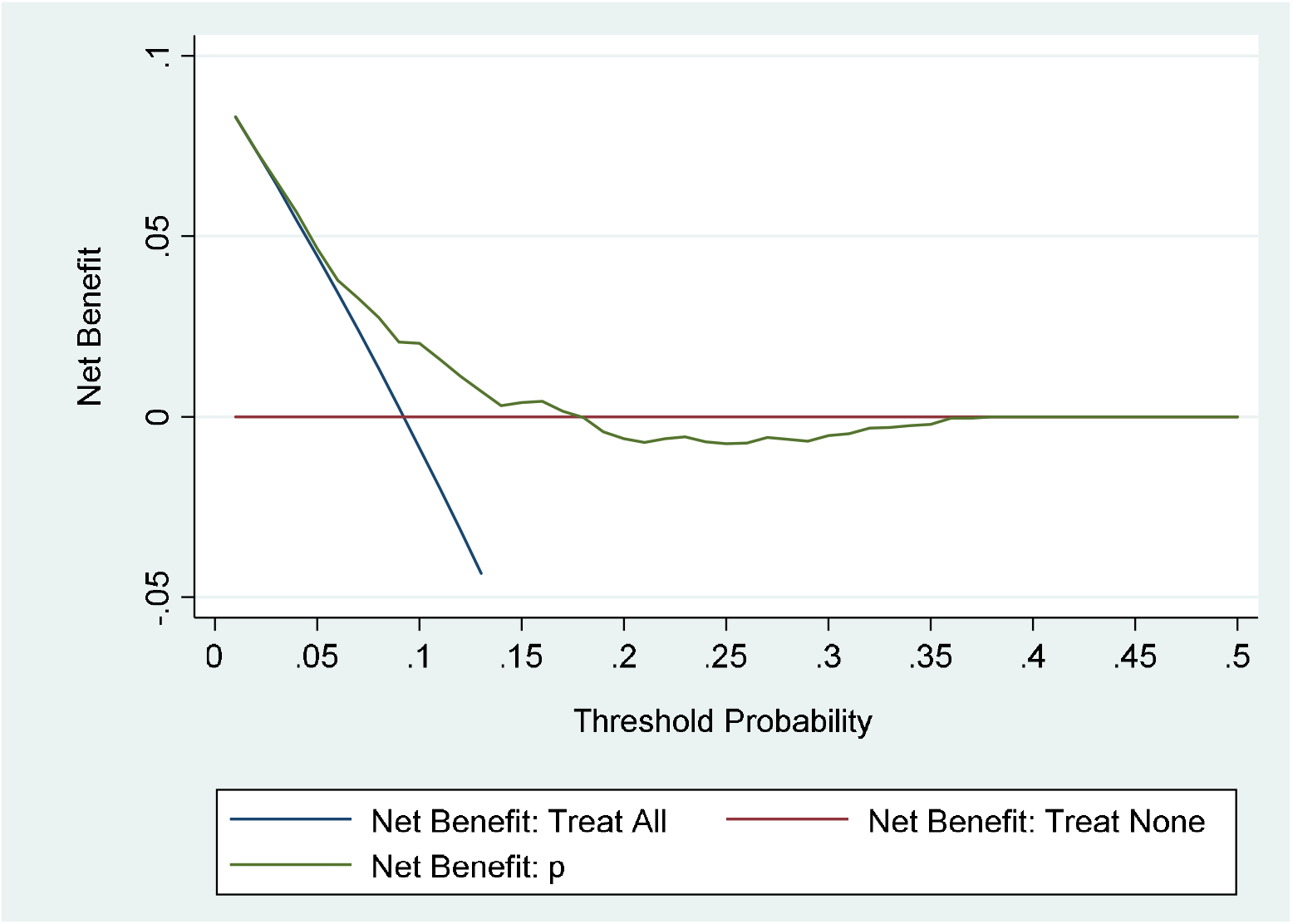
Decision curve analysis in the validation data sets.

## Discussion

We assessed the predictive performance of the diagnostic model in the validation data sets by examining measures of discrimination, calibration and DCA. AUC was 0.6547±0.025,95% CI= 0.605∼0.704.Brier score = 0.0997<0.25.When P is less than 17%, the clinical value is high; the prediction formula or nomogram can be used normally. When P is greater than or equal to 17%, the clinical value is poor; the prediction formula or nomogram is not suitable for clinical use.Discrimination, calibration, and DCA were satisfactory.

We investigated the predisposing factors of CMVO/NR in patients with acute STEMI during PPCI. A frequency of CMVO/NR was 11.9%(147/1232). Age, periprocedural bradycardiais, using thrombus aspiration devices during procedure,and total occlusion of culprit vessel were independent risk factors of CMVO/NR. We can use the formula or nomogram to predicte CMVO/NR.

We followed the discussion of Li et al.^[13]^

### Study Limitations

This was a single center experience. Some patients were enrolled >10 years ago thus their treatment may not conform to current standards and techniques. We wanted to get risk factors of CMVO/NR before it happen, some variables associated with it is not including, so the c statistic of the model at 0.6833 in the derivation and 0.6547 in the validation cohort was modest.

## Conclusions

We developed and externally validated a diagnostic model of CMVO/NR during PPCI.

## Data Availability

Data availability statement
The data used to support the findings of this study are included within the supplementary material.
The data are demographic, clinical, and angiographic characteristics of patients with acute STEMI during PPCI. CNR= CMVO/NR;AGE= age; S = sex; HH = history of hypertension; HD= history of diabetes; MIH=history of myocardial infarction;AH= History of angina; PCIH=history of percutaneous coronary intervention ; PB=periprocedural bradycardia; IH = intra-procedural hypotension ; TOCV=Total occlusion of the culprit vessel; RCA= the culprit vessel was right coronary artery ;LAD= the culprit vessel was left anterior descending coronary artery;LCX= the culprit vessel was left circumflex coronary artery.

https://pan.baidu.com/s/1HpbSBXMxsUK96NGPS-qlYQ

## Declarations

### Compliance with ethical Standards

The study was approved by ethics committee. Approved No. of ethic committee: 2019013X. Name of the ethics committee:Ethics Committee of Beijing Anzhen Hospital Capital Medical University. It was a retrospective analysis and informed consent was waived by Ethics Committee of Beijing Anzhen Hospital Capital Medical University.

### Statement of human and animal rights

All procedures performed in studies involving human participants were in accordance with the ethical standards of the institutional and/or national research committee and with the 1964 Helsinki declaration and its later amendments or comparable ethical standards. The study was not conducted with animals.

### Contributor ship statement

Yong Li contributed to generating, analysing, and interpreting the study data and drafted the manuscript. Shuzheng Lyu contributed what to the planning and revised the manuscript critically for important intellectual content. All authors reviewed the manuscript.

Yong Li and Shuzheng Lyu are being responsible for the overall content as guarantor.

### Data availability statement

The data used to support the findings of this study are included within the supplementary material. The data are demographic, clinical, and angiographic characteristics of patients with acute STEMI during PPCI. CNR= CMVO/NR;AGE= age; S = sex; HH = history of hypertension; HD= history of diabetes; MIH=history of myocardial infarction;AH= History of angina; PCIH=history of percutaneous coronary intervention ; PB=periprocedural bradycardia; IH = intra-procedural hypotension ; TOCV=Total occlusion of the culprit vessel; RCA= the culprit vessel was right coronary artery ;LAD= the culprit vessel was left anterior descending coronary artery;LCX= the culprit vessel was left circumflex coronary artery.

## References

[1] Bouleti C, Mewton N, Germain S. The no-reflow phenomenon: State of the art. Arch Cardiovasc Dis 2015;108:661–74.

[2] Rezkalla SH, Stankowski RV, Hanna J, Kloner RA. Management of No-Reflow Phenomenon in the Catheterization Laboratory. JACC Cardiovasc Interv 2017;10:215–23.

[3] Karimianpour A, Maran A. Advances in Coronary No-Reflow Phenomenon-a Contemporary Review. Curr Atheroscler Rep 2018;20:44.

[4] Durante A. Role of no reflow and microvascular obstruction in the prognostic stratification of STEMI patients. Anatol J Cardiol, 2018,19(5):346–349.

[5] Li Y, Lyu S. Risk Factors of Periprocedural Bradycardia during Primary Percutaneous Coronary Intervention in Patients with Acute ST-Elevation Myocardial Infarction. Cardiol Res Pract. 2019. 2019: 4184702.

[6] Yong Li. Development and External Validation of a Diagnostic Model for in-Hospital Mortality in Patient with Acute ST Elevation Myocardial Infarction. medRxiv 2020.05.28.20115485; doi: https://doi.org/10.1101/2020.05.28.20115485.

[7] Moons KG, Altman DG, Reitsma JB, et al. Transparent Reporting of a multivariable prediction model for Individual Prognosis or Diagnosis (TRIPOD): explanation and elaboration. Ann Intern Med 2015;162:W1–73.

[8] Thygesen K, Alpert JS, Jaffe AS, et al. Fourth Universal Definition of Myocardial Infarction (2018). J Am Coll Cardiol. 2018. 72(18): 2231–2264.

[9] Ibanez B, James S, Agewall S, et al. 2017 ESC Guidelines for the management of acute myocardial infarction in patients presenting with ST-segment elevation: The Task Force for the management of acute myocardial infarction in patients presenting with ST-segment elevation of the European Society of Cardiology (ESC). Eur Heart J, 2018,39(2):119–177.

[10] Niccoli G, Scalone G, Lerman A, Crea F. Coronary microvascular obstruction in acute myocardial infarction. Eur Heart J 2016;37:1024–33.

[11] Kusumoto FM, Schoenfeld MH, Barrett C, et al. 2018 ACC/AHA/HRS Guideline on the Evaluation and Management of Patients With Bradycardia and Cardiac Conduction Delay: A Report of the American College of Cardiology/American Heart Association Task Force on Clinical Practice Guidelines and the Heart Rhythm Society. Circulation. 2019. 140(8): e382–e482.

[12] Baran DA, Grines CL, Bailey S, et al. SCAI clinical expert consensus statement on the classification of cardiogenic shock: This document was endorsed by the American College of Cardiology (ACC), the American Heart Association (AHA), the Society of Critical Care Medicine (SCCM), and the Society of Thoracic Surgeons (STS) in April 2019. Catheter Cardiovasc Interv. 2019. 94(1): 29–37.

[13] Li, Yong Lyu, Shuzheng Risk factors of coronary microvascular obstruction MedRxiv 2020.05.29.20116665 DOI: 10.1101/2020.05.29.20116665.

